# Deep Learning-enabled Detection of Aortic Stenosis from Noisy Single Lead Electrocardiograms

**DOI:** 10.1101/2023.09.29.23296310

**Authors:** Arya Aminorroaya, Lovedeep S Dhingra, Veer Sangha, Evangelos K Oikonomou, Akshay Khunte, Sumukh Vasisht Shankar, Aline Pedroso Camargos, Norrisa A Haynes, Ira Hofer, David Ouyang, Girish N. Nadkarni, Rohan Khera

## Abstract

**Background:** Due to the lack of a feasible screening strategy, aortic stenosis (AS) is often diagnosed after the development of clinical symptoms, representing advanced stages of disease. Portable and wearable devices capable of recording electrocardiograms (ECGs) can be used for scalable screening for AS, if the diagnosis can be made with a single-lead ECG, despite potentially noisy acquisition.

**Methods:** Using electronic health records and imaging data from a large, diverse hospital system (2015-2022), we developed a deep learning-based approach to detect moderate/severe AS using a single-lead ECG. We used ECGs paired with echocardiograms obtained within 30 days of each other to develop the model. We extracted lead I signal data from clinical ECG and augmented it with random Gaussian noise. We trained a convolutional neural network (CNN) to identify TTE-confirmed AS using noisy single-lead ECGs. Finally, we used the CNN model probabilities, along with patient age and sex, as predictive inputs to train an extreme gradient boosting (XGBoost) model to detect moderate/severe AS.

**Results:** The model was developed in 75,901 ECGs/35,992 patients (median age 61 [interquartile range (IQR) 47-72] years, 54.3% women, 9.5% Black) and validated in 3,733 patients (median age 61 [IQR 47-72] years, 53.4% women, 9.7% Black). In the held-out validation set, the ensemble XGBoost model achieved an AUROC of 0.829 (95% CI: 0.800-0.855), with a sensitivity of 90.4% and specificity of 58.7% for detecting moderate/severe AS. For detecting severe AS, the model’s AUROC was 0.846 (95% CI, 0.778-0.899), with a sensitivity of 94.3% and specificity of 57.0%. In the test set with a 4.5% prevalence of moderate/severe AS, the model had a PPV of 9.3% and an NPV of 99.2%. In simulated cohorts with 1% and 20% prevalence of moderate/severe AS, the model’s NPVs varied from 99.8% to 96.1%, and PPV from 2.2% to 35.4%, respectively.

**Conclusion:** We developed a novel portable– and wearable-adapted deep learning approach for the detection of moderate/severe AS from noisy single-lead ECGs. Our approach represents a highly sensitive, feasible, and scalable strategy for community-based AS screening.

## BACKGROUND

Aortic stenosis (AS) poses a significant burden of cardiovascular morbidity and mortality that is potentially treatable, but the lack of a feasible strategy for early detection of disease challenges the identification of individuals before the onset of symptoms. Prevalence of AS increases with advancing age, with a prevalence of 10-15% among individuals over 70 years of age.^1–4^ The clinical progression of AS is associated with worsening obstruction of left ventricular outflow, an associated decrease in exercise capacity, and the increased risk of exertional angina, syncope, heart failure, and sudden cardiac death.^5^ Furthermore, severe AS poses a substantial burden on health systems globally, with substantial quality-adjusted life years (QALY) lost as well as nearly $30,000, per patient, in healthcare expenses attributable to AS annually.^6^ This is despite the presence of effective treatment with aortic valve replacement (AVR), including with a low-risk transcatheter approach (TAVR).^7,8^ Moreover, AS is often diagnosed after the development of clinical symptoms, representing advanced stages of disease.^9,10^ This is because the diagnosis of AS relies on echocardiographic Doppler imaging, which requires access to specialized imaging equipment and trained personnel, limiting its scalability.

While echocardiography remains impractical as a screening strategy, the application of artificial intelligence algorithms on ECGs (AI-ECG) has been proposed for the automated detection of AS.^11–14^ However, the current AI-ECG approaches rely on the use of 12-lead clinical ECGs, which do not allow their use as a community-based screening modality. Using portable and wearable devices capable of capturing single lead ECGs can facilitate large-scale cardiovascular screening.^15–17^ However, these devices are often prone to the introduction of noise during ECG acquisition.^16,18^ Thus, the advancement of scalable strategies for AS detection requires development of noise-resilient AI-ECG models capable of detecting AS from single-lead ECGs.

In this study, we use data from a diverse patient population for the development and validation of a deep learning-enabled strategy for the screening of AS using noisy single-lead ECGs.

## METHODS

The Yale Institutional Review Board approved the study protocol and waived the need for informed consent as the study involves secondary analysis of pre-existing data.

### Data Source and Study Population

We used voltage data for lead I extracted from 12-lead ECGs obtained at the Yale New Haven Hospital (YNHH) during 2015-2022. These ECGs were acquired during routine clinical practice as standard 10-second 12-lead ECGs at a sampling frequency of 500 Hz. We included ECGs from patients who received a transthoracic echocardiogram (TTE) within 30 days before or after obtaining the ECG. TTEs were conducted by cardiac sonographers and interpreted by expert cardiologists at the YNHH. We utilized TTEs conducted in an outpatient setting and encompassing a complete structural assessment of the heart, given that inpatient TTEs are more likely to be affected by a patient’s evolving condition during hospitalization.

Moreover, we excluded TTEs that were conducted after undergoing cardiac surgical procedures, such as coronary artery bypass grafting, valve replacement procedures, left ventricular assist device implantation, heart transplant, alcohol septal ablation, and ventricular myectomy, or for acute indications, including myocardial infarction, stroke, pulmonary edema, and decompensated heart failure. This ensures the model learns the pathological ECG signatures relevant to detecting AS in a population without prior cardiac assessment. Demographics, ECG, and TTE data were extracted from the YNHH electronic health record.

### Study Outcome

The severity of AS was graded by the reading expert cardiologist as sclerosis without stenosis, mild, moderate, and severe stenosis according to the European Association of Cardiovascular Imaging and the American Society of Echocardiography recommendations.^19^ Briefly, a peak aortic velocity of ≥4.0 m/s, a mean gradient of ≥40 mmHg, or an aortic valve area of <1 cm^2^ were considered as severe AS, while mild AS was defined as a peak aortic velocity between 2.6 and 2.9 m/s, a mean gradient of <20 mmHg, or an aortic valve area of >1.5 cm^2^. Moderate AS was identified when echocardiographic measures did not meet the criteria for mild or severe cases. Aortic sclerosis without stenosis was characterized by a peak aortic velocity of ≤2.5 m/s, despite presence of aortic sclerosis. We defined any moderate or severe AS as the primary study outcome. We also reported the model performance for detecting severe AS.

### Signal Preprocessing

We extracted the signal data from lead I of 12-lead ECG recordings, which represents the standard lead captured by wearable devices. Our signal preprocessing strategy involved median pass filtering, scaling, normalizing, and winsorizing for each lead. Median filtering was conducted by subtracting a one-second median filter from the acquired signals to eliminate baseline drift. ECG signals were divided by 1000 to scale them to millivolts. Next, we normalized every signal data point by computing the difference between the data point and the mean of voltage signals, divided by the standard deviation of voltage signals in each lead. We then winsorized voltage values by removing values that fell outside the 1^st^ or 99^th^ percentile in each lead. Ultimately, we isolated preprocessed signal data from lead I ECG.

### Noising Strategy

To construct algorithms resilient to the noise introduced during acquisition of ECGs obtained from wearable and portable devices, we artificially incorporated noises into the model development process, as described previously.^16^ Briefly, we augmented ECGs in the training set using random Gaussian noises, while we tested the model on clean ECGs without incorporating noises. We isolated 4 distinct noises from a 5-minute random Gaussian noise within 4 frequency ranges of 3-12 Hz, 12-50 Hz, 50-100 Hz, and 100-150 Hz, each corresponding to the frequency range of a specific type of real-world noise. Each ECG in the training set was included 2 times with different random noises in training the model. This augmentation involved a random type of noise and a random signal-to-noise ratio (SNR). For this purpose, we first randomly selected 1 of the 4 abovementioned distinct random Gaussian noises. Finally, the selected noise was introduced to the ECG waveform with a random SNR ranging from 0.5 to 1.25, representing a heavy and a light burden of noise in ECGs, respectively.

### Model Development

We randomly split the included ECG-TTE pairs into training, validation, and test sets with a ratio of 85:5:10 at the patient level. To allow for the broadest training set, we used all ECG-TTE pairs for a patient in the training set. The validation and test sets included only one random ECG per patient. We used transfer learning to train a CNN model to detect the presence of moderate/severe AS on noisy single-lead ECGs. Next, we trained an extreme gradient boosting (XGBoost) classifier model using the CNN model probabilities and the patient’s age and sex as predictive features.

For transfer learning, we first trained a CNN model to detect left ventricular systolic dysfunction (LVSD; defined as left ventricular ejection fraction < 40%). Subsequently, weights from the LVSD model were used to initialize the training for the model for AS detection. We evaluated multiple convolutional neural network (CNN) architectures, experimenting with the number and size of convolutional layers as well as dropout and learning rate. The architecture with the highest area under the receiver operating characteristic curve (AUROC) for detecting LVSD – a commonly hidden label on ECGs – in the validation set was selected as the final architecture for training. This final architecture comprised an input layer with dimensions of (5000, 1, 1), representing a 10-second, 500 Hz, lead I ECG. The input layer was followed by seven 2-dimensional convolutional layers, progressively increasing the number of filters from 16 to 64 while incorporating varying kernel sizes (7×1, 5×1, and 3×1) to capture different levels of feature abstraction. A batch normalization layer, a ReLU activation layer, and a 2-dimensional max-pooling layer with different pool sizes (2×1 and 4×1) followed each convolutional layer. Next, the output of the 7^th^ convolutional layer was used as the input for a fully connected network that included two dense layers. Each dense layer was followed by a batch normalization layer, a ReLU activation layer, and a dropout layer with a rate of 0.5. Finally, the model output was a dense layer with a single class and a sigmoid activation to generate the output probability of the label. The loss function was adjusted by calculating model weights using the effective number of samples class re-weighting approach to ensure that the learning is not impacted by the differential prevalence of positive and negative labels.^16^

The model training was stopped when the validation loss did not improve for 10 consecutive epochs. After the completion of model training, the optimal epoch was selected based on low validation loss and high validation area under the receiver operating characteristic curve (AUROC).

For training the XGBoost model, patient age and the CNN model output probabilities were standard scaled to achieve a mean of 0 and a variance of 1 before being included as features in the model. The model hyperparameters were finetuned using Scikit-learn’s GridSearchCV. Then, the XGBoost model performance was evaluated in the test set. Finally, we evaluated SHapley Additive exPlanations (SHAP) values in the held-out test set to explain the interpretability of the final ensemble model.^20^

### Statistical Analysis

Continuous variables were reported as mean (standard deviation [SD]) or median (interquartile range [IQR]), as appropriate, and categorical variables as number (percentage%). The model’s performance was presented as AUROC and area under the precision-recall curve (AUPRC). Furthermore, we reported sensitivity, specificity, positive predictive value (PPV), negative predictive value (NPV), and F1 score of the model across the thresholds and specifically for the threshold for optimizing sensitivity at 90%. The 95% confidence intervals (CI) for AUROC and AUPRC were calculated using bootstrapping with 1000 iterations. We computed 95% CI for sensitivity, specificity, PPV, NPV, and F1 score using the standard error formula for proportion. Finally, we calculated the model’s PPV in simulated screening scenarios with varying prevalence of moderate/severe AS, and severe AS using the threshold for optimizing sensitivity at 90%. The statistical significance level was set at P≤0.05. All statistical analyses were executed using Python 3.11.2, and R version 4.2.0.

## RESULTS

### Study Population

From September 2015 to January 2022, we included 528,603 ECGs from 131,804 patients with a TTE obtained within a 30-day window of the ECG at YNHH. After retaining TTEs with a complete assessment of the aortic valve and applying other eligibility criteria, we included 75,901 ECG-TTE pairs, representing 35,992 unique patients, for the model development.

The study population had a median age of 61 (IQR, 47-72) years and included 19,488 (54.3%) women, 3,276 (9.5%) of non-Hispanic Black race, 2,922 (8.5%) of Hispanic ethnicity, and 742 (2.2%) of Asian race. There were 5,782 (7.6%) ECG-TTE pairs with moderate/severe AS, including 4,056 (5.3%) with moderate AS and 1,726 (2.3%) with severe AS, representing 1,544 (4.3%), 1,189 (3.3%), and 355 (1.0%) unique patients (**Table 1**). The training set comprised 30,403 patients with 64,343 ECG-TTE pairs. The validation and held-out test sets included 1,856 patients and 3,733 patients, respectively, with one ECG per patient.

**Table 1.** Demographics and Prevalence of Aortic Stenosis in the Development Set at Patient– and ECG-level. Abbreviations: AS, aortic stenosis; ECG, electrocardiogram.

| **Characteristic\*** | **Patients** | **ECGs** |
| --- | --- | --- |
| **Number** | 35,992 | 75,901 |
| **Demographics** |  |  |
| **Age (years)** | 61 [47-72] | 64 [52-74] |
| **Female Sex** | 19,488 (54.3%) | 37,828 (49.9%) |
| **Race and Ethnicity** |  |  |
| White | 27,055 (78.7%) | 57,419 (78.5%) |
| Black | 3,276 (9.5%) | 7,790 (10.7%) |
| Hispanic | 2,922 (8.5%) | 5,830 (8.0%) |
| Asian | 742 (2.2%) | 1,290 (1.8%) |
| Other | 371 (1.1%) | 776 (1.1%) |
| **Moderate AS** | 1,189 (3.3%) | 4,056 (5.3%) |
| **Severe AS** | 355 (1.0%) | 1,726 (2.3%) |
| **Moderate/Severe AS** | 1,544 (4.3%) | 5,782 (7.6%) |

\*Data are presented as median [interquartile range], or number (percentage%).

### Detection of Moderate/Severe AS, and Severe AS

The 1-lead ECG-only CNN model achieved an AUROC of 0.755 (95% CI, 0.722-0.789) and an AUPRC of 0.126 (95% CI, 0.101-0.164) for detecting moderate/severe AS with a prevalence of 4.5% in the held-out test (**Figure 2**). For detecting severe AS, the 1-lead ECG-only CNN model demonstrated an AUROC of 0.809 (95% CI, 0.726-0.889) and an AUPRC of 0.065 (95% CI, 0.035-0.136) with a prevalence of severe AS of 0.9% in the test set. The composite XGBoost model that included age and sex achieved an AUROC of 0.829 (95% CI, 0.800-0.855) and an AUPRC of 0.166 (95% CI, 0.134-0.215) for detecting moderate/severe AS, and an AUROC of 0.846 (95% CI, 0.778-0.899) and an AUPRC of 0.068 (95% CI, 0.032-0.144) for detecting severe AS (**Figure 2**).

**Figure 1.**
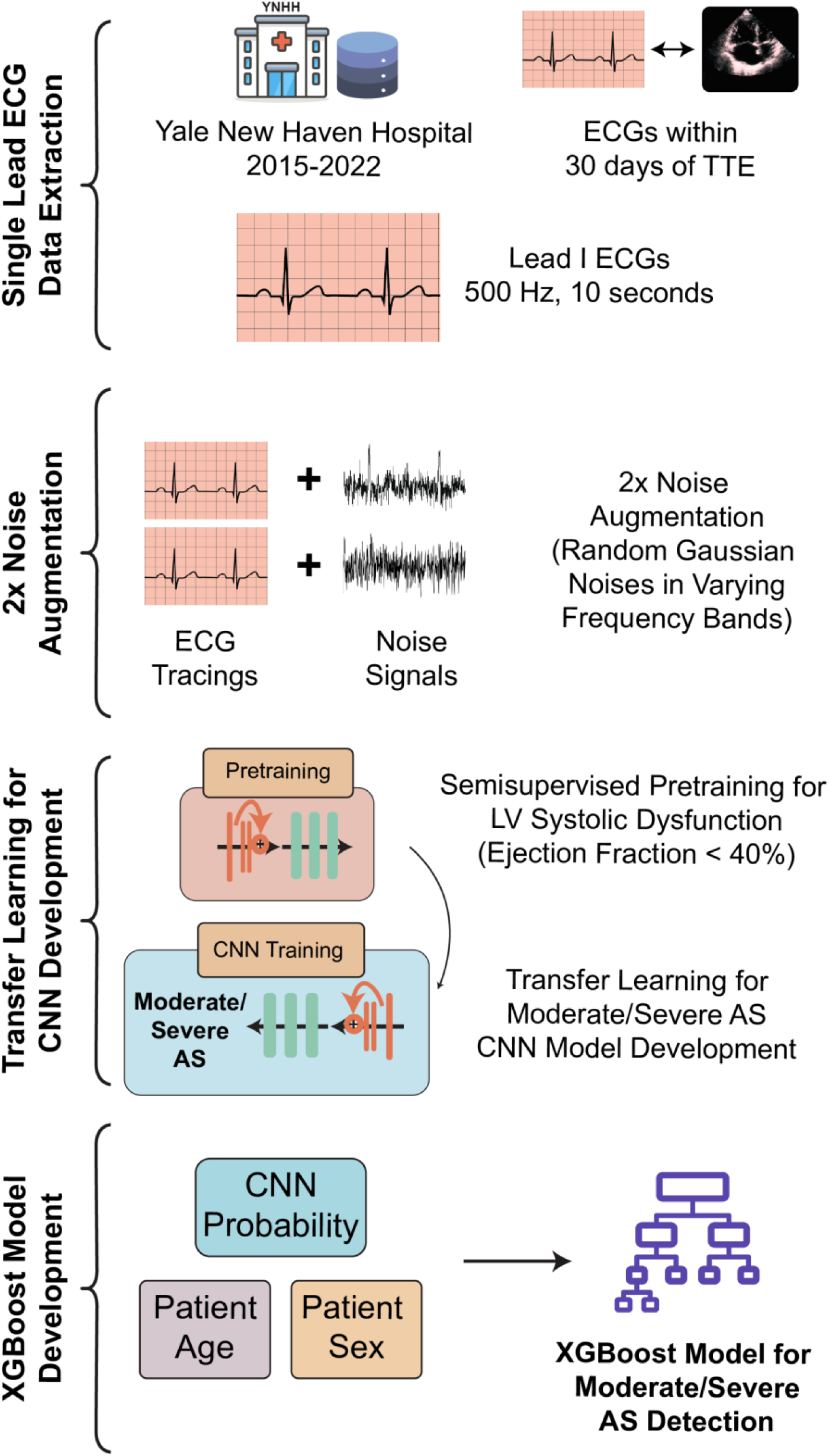
Model Development for Detecting Multiple Structural Heart Diseases. Abbreviations: AS, aortic stenosis; CNN, convolutional neural network; ECG, electrocardiogram; XGBoost, extreme gradient boosting.

**Figure 2.**
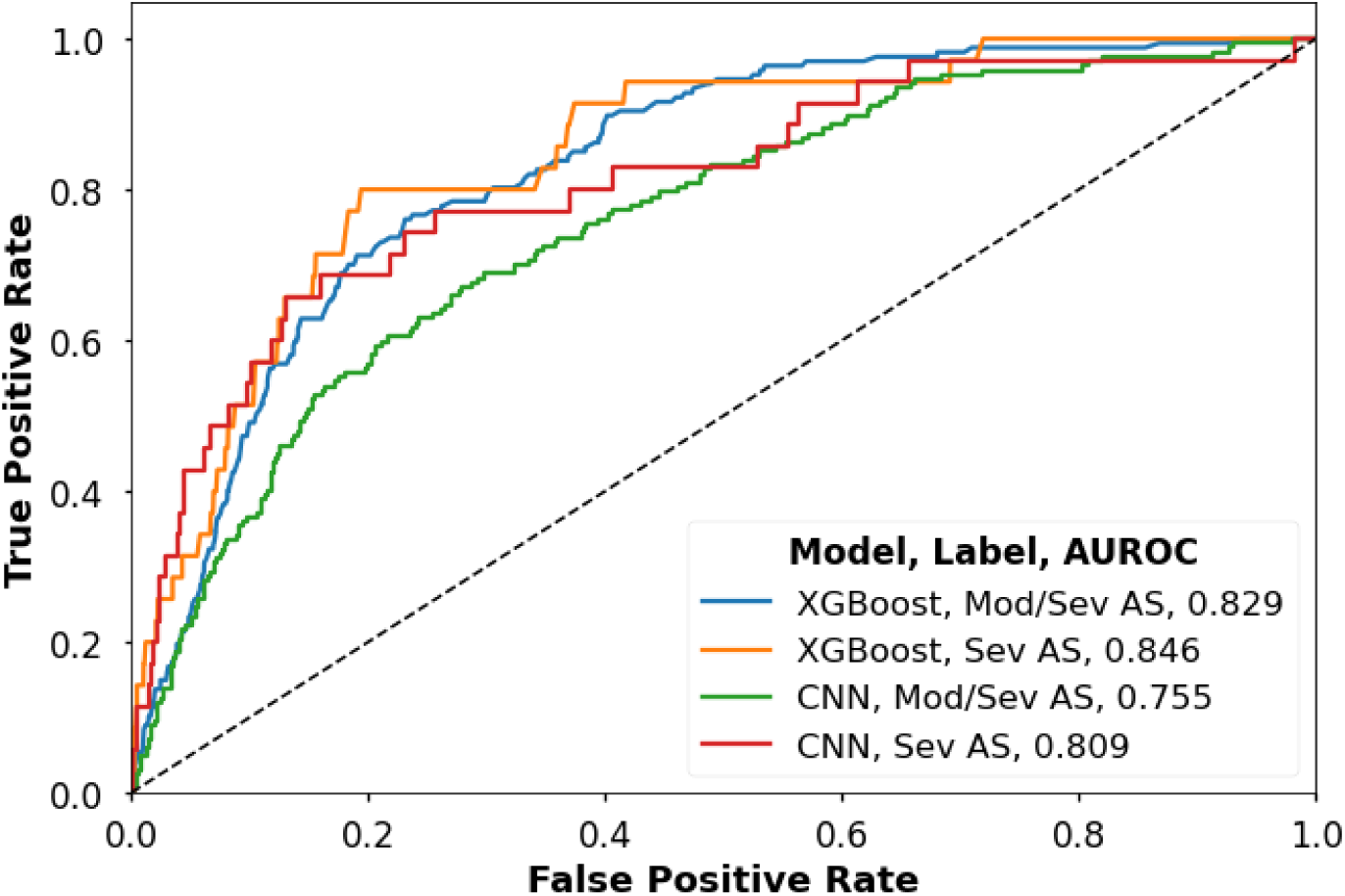
Receiver Operating Characteristic Curves for Detecting Moderate/Severe Aortic Stenosis, and Severe Aortic Stenosis Across Model Architectures. Abbreviations: AS, aortic stenosis; AUROC, area under the receiver operating characteristic curve; CNN, convolutional neural network; Mod/Sev, moderate/severe; Sev, severe; XGBoost, extreme gradient boosting.

The highest F1 score and Youden’s index of the XGBoost model were 0.277 and 0.529, respectively, in the held-out test set. At the threshold for optimizing sensitivity at 90%, and at a prevalence of 4.5%, the model showed a sensitivity of 0.904 (95% CI, 0.895-0.914), specificity of 0.587 (95% CI, 0.571-0.603), PPV of 0.093 (95% CI, 0.084-0.102), and NPV of 0.992 (0.990-0.995) for detecting moderate/severe AS (**Table 2**, **Figure 3**). For detecting severe AS, at the same threshold, the model had a sensitivity of 0.943 (95% CI, 0.935-0.950), specificity of 0.570 (95% CI, 0.554-0.586), PPV of 0.020 (95% CI, 0.016-0.025), and NPV of 0.999 (95% CI, 0.998-1.000). Using the threshold for optimizing both sensitivity and specificity, the model achieved a sensitivity of 0.760 and a specificity of 0.769 for detecting moderate/severe AS, and a sensitivity of 0.800 and a specificity of 0.750 for detecting severe AS. At the threshold for optimizing the F1 score, the model demonstrated a sensitivity of 0.563 and a specificity of 0.883 for detecting moderate/severe AS, and a sensitivity of 0.657 and a specificity of 0.868 for detecting severe AS.

**Figure 3.**
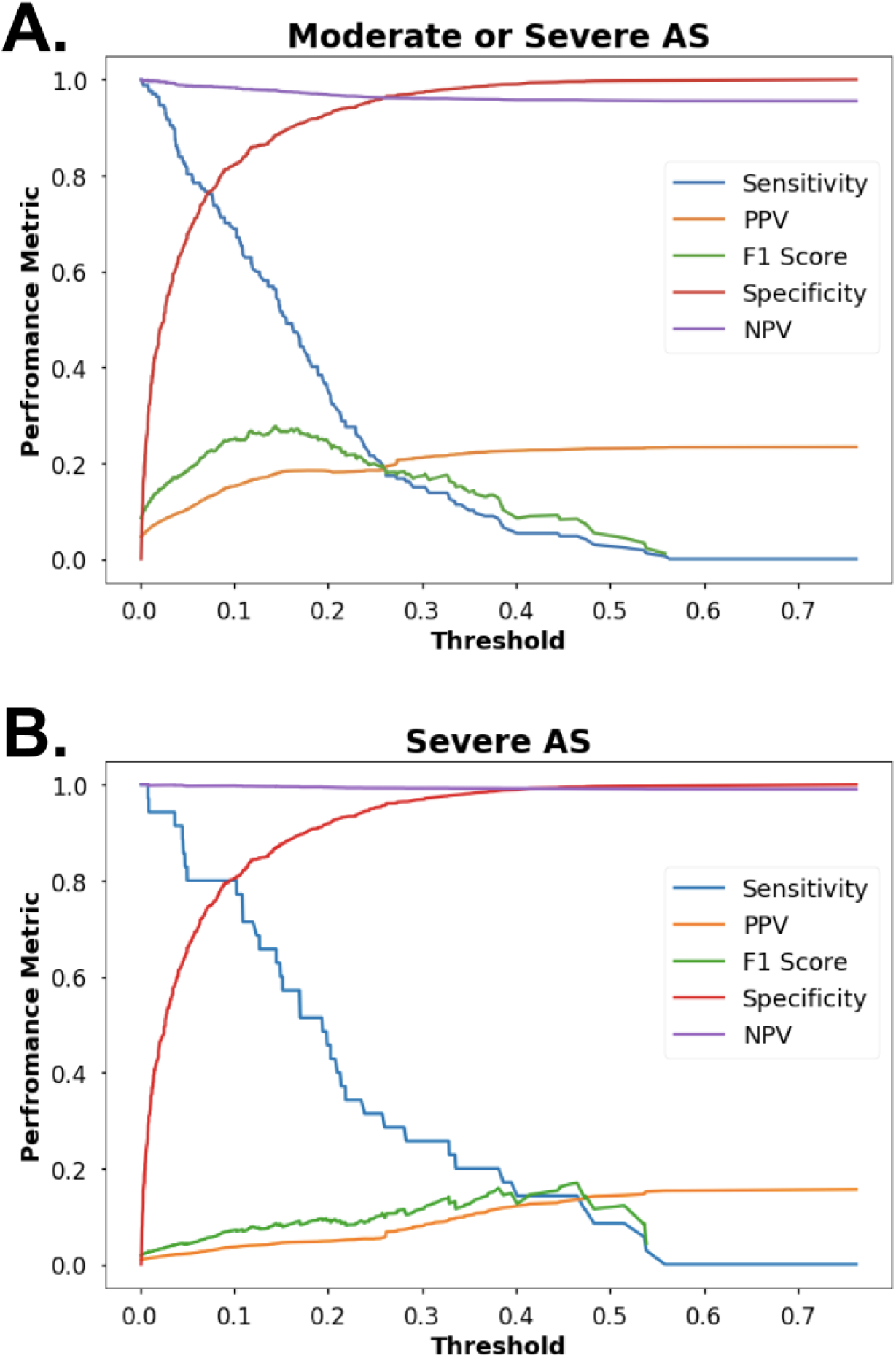
Model Performance Measures Across Thresholds for Detecting (A) Moderate/Severe Aortic Stenosis, and (B) Severe Aortic Stenosis. Abbreviations: AS, aortic stenosis; NPV, negative predictive value; PPV, positive predictive value.

**Table 2.** Performance Measures of the Model for Detecting Moderate/Severe Aortic Stenosis and Severe Aortic Stenosis Across Thresholds in the Held-out Test Set. Abbreviations: AS, aortic stenosis; NPV, negative predictive value; PPV, positive predictive value.

| **Threshold** | **Sensitivity**<br>**(95% CI)** | **Specificity**<br>**(95% CI)** | **PPV (95% CI)** | **NPV (95% CI)** |
| --- | --- | --- | --- | --- |
| **Moderate/Severe AS** |  |  |  |  |
| 90% Sensitivity | 0.904 (0.895-0.914) | 0.587 (0.571-0.603) | 0.093 (0.084-0.102) | 0.992 (0.990-0.995) |
| Optimal Youden Index | 0.760 (0.747-0.774) | 0.769 (0.755-0.782) | 0.133 (0.122-0.144) | 0.986 (0.982-0.989) |
| Optimal F1-score | 0.563 (0.547-0.579) | 0.883 (0.872-0.893) | 0.184 (0.171-0.196) | 0.977 (0.973-0.982) |
| 90% Specificity | 0.491 (0.475-0.507) | 0.900 (0.891-0.910) | 0.187 (0.175-0.200) | 0.974 (0.969-0.979) |
| **Severe AS** |  |  |  |  |
| 90% Sensitivity | 0.943 (0.935-0.950) | 0.570 (0.554-0.586) | 0.020 (0.016-0.025) | 0.999 (0.998-1.000) |
| Optimal Youden Index | 0.800 (0.787-0.813) | 0.750 (0.736-0.764) | 0.029 (0.024-0.035) | 0.997 (0.996-0.999) |
| Optimal F1-score | 0.657 (0.642-0.672) | 0.868 (0.857-0.879) | 0.045 (0.038-0.052) | 0.996 (0.994-0.998) |
| 90% Specificity | 0.571 (0.556-0.587) | 0.887 (0.877-0.897) | 0.046 (0.039-0.052) | 0.995 (0.993-0.998) |

### Simulated Screening Scenarios

In simulated screening scenarios, at the threshold for optimizing sensitivity, the model achieved a PPV ranging from 2.2% to 35.4% for detecting moderate/severe AS considering a prevalence of 1% to 20% in simulated populations, respectively (**Table 3**). For severe AS, we considered simulated populations with a prevalence ranging from 0.1% to 10%, where the model demonstrated a PPV ranging from 0.2% to 19.6%, respectively. The NPV was greater than 99% in all scenarios for detecting severe AS.

**Table 3.**
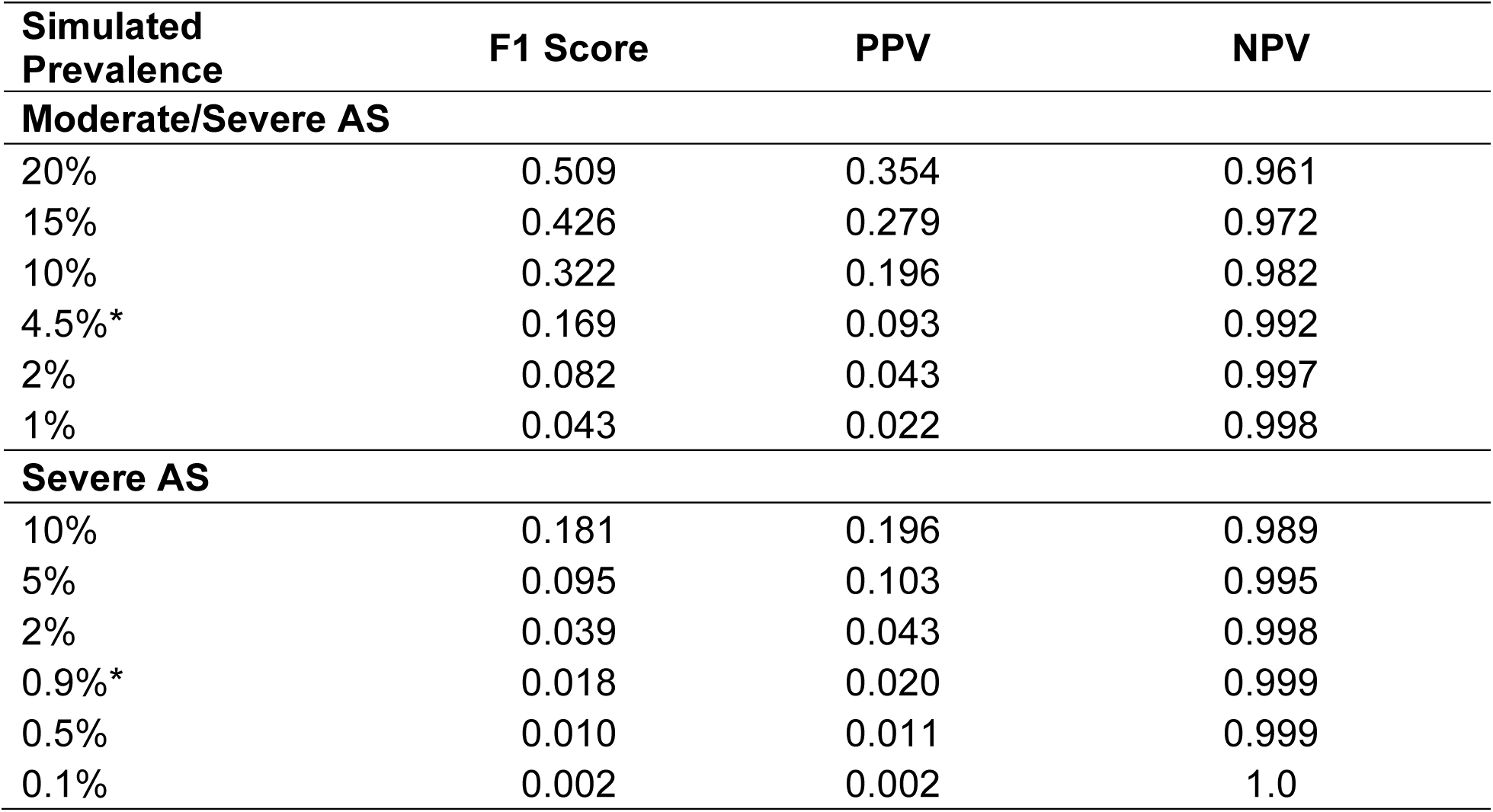
Performance Metrics for Detecting Moderate/Severe Aortic Stenosis, and Severe Aortic Stenosis in Simulated Screening Scenarios at the Threshold for 90% Sensitivity. Abbreviations: AS, aortic stenosis; NPV, negative predictive value; PPV, positive predictive value. *Prevalence of SHD in the held-out test set.

| **Simulated Prevalence** | **F1 Score** | **PPV** | **NPV** |
| --- | --- | --- | --- |
| **Moderate/Severe AS** |  |  |  |
| 20% | 0.509 | 0.354 | 0.961 |
| 15% | 0.426 | 0.279 | 0.972 |
| 10% | 0.322 | 0.196 | 0.982 |
| 4.5%* | 0.169 | 0.093 | 0.992 |
| 2% | 0.082 | 0.043 | 0.997 |
| 1% | 0.043 | 0.022 | 0.998 |
| **Severe AS** |  |  |  |
| 10% | 0.181 | 0.196 | 0.989 |
| 5% | 0.095 | 0.103 | 0.995 |
| 2% | 0.039 | 0.043 | 0.998 |
| 0.9%* | 0.018 | 0.020 | 0.999 |
| 0.5% | 0.010 | 0.011 | 0.999 |
| 0.1% | 0.002 | 0.002 | 1.0 |

## DISCUSSION

We developed a noise-adapted deep learning strategy for detecting advanced AS from noisy single lead ECG using data from a diverse population at a large US health system. The model achieved a high sensitivity of greater than 90% and modest specificity of 59% for detecting moderate/severe AS using single-lead portable ECG signals, accounting for their real-world noisy acquisition. The approach may represent an ideal first-stage community-based screening for moderate/severe AS, where the low prevalence of the condition makes echocardiographic screening infeasible.

Our study expands on the existing literature on using deep learning for electrocardiographic detection of advanced AS in two significant ways. First, we introduce a model capable of detecting moderate/severe AS from single-lead ECGs, adapting to their quality obtainable on portable and wearable devices. Second, our model development approach addresses the challenge of noisy ECG acquisition using wearable and portable devices. Previous studies have developed AI models to detect severe AS using clinical 12-lead ECGs, demonstrating AUROCs ranging from 0.87-0.91^11–14^ However, our algorithm achieves an AUROC of 0.85 for detecting severe AS, while relying solely on data from lead I ECG. Consequently, our model’s proposed use on portable device-acquired ECGs represents an accessible approach for detecting moderate/severe AS in the community, as these portable devices are widely available, and easily deployable.^15,21^ Finally, our model incorporates real-world noise during the model training, which enhances the model’s ability to identify the pathological disease signature even in the presence of noise in portable ECG acquisition.^13,16^

The morbidity and mortality arising from AS is treatable with recent advances in transcatheter aortic valve replacement, making AS management safer and more feasible.^22–24^ However, the diagnosis of AS is frequently missed in routine care, with only 10 to 20% of people with moderate/severe AS in the community being identified.^25–27^ Despite the highlighted importance of AS screening, widespread use of TTE does not represent a feasible screening strategy for AS.^16,28–31^ Thus, the application of deep learning on ECGs (AI-ECG) and point-of-care ultrasonographic images (AI-POCUS) have been proposed as modalities for the detection of AS.^32^ While AI-POCUS represents an easy-to-operate and accurate strategy for automated AS detection, its deployment in a community setting may still be limited by the access to handheld echocardiographic devices. Alternatively, the application of our model using portable ECG devices can represent a highly scalable and sensitive strategy for finding people with undiagnosed AS in the community. This screening test can be followed by AI-POCUS or TTE for screen-positive people to detect AS, particularly among older adults who are at higher risk of AS.^1^

Our study has several limitations. First, the development set’s composition may not fully represent a real-world screening population. The study population underwent TTE in close proximity to their ECG acquisition, which is not necessarily reflective of routine clinical practice where TTE might be conducted at varying time intervals. However, since model development in a screening population setting is impractical, and would require protocolized TTE to be performed in a large cohort, it is essential to prospectively validate the current strategy before widespread deployment. Second, our model was developed using data from a single tertiary health system. However, YNHH serves a diverse population with a catchment area spanning across multiple states. Moreover, the New Haven county is one of the most representative of the adult US population,^33^ thus enabling model generalizability across a wide population. Third, we trained our model using lead I ECG data extracted from 12-lead ECGs, as portable or wearable ECGs paired with TTE are not available for model development. This might limit the model’s applicability to scenarios involving real-world portable and wearable ECG devices. However, we augmented single-lead ECGs with random Gaussian noises during the model training to enable identification of pathological electrical signature from noisy ECGs, that could represent real-world acquisition.

## CONCLUSION

We developed a highly sensitive noise-adapted AI-ECG algorithm that detects moderate/severe AS using single-lead ECG acquired by a portable device. The model holds the potential for AS screening to address the undiagnosed, yet treatable burden of AS in the community.

## Data Availability

The data used represent protected health information. Thus, they are not available for public use.

